# The Impact of Income Support Systems on Healthcare Quality and Functional Capacity in Workers with Low Back Pain: A Realist Review

**DOI:** 10.1101/2020.01.13.20017343

**Authors:** Michael Di Donato, Ross Iles, Tyler Lane, Rachelle Buchbinder, Alex Collie

## Abstract

**Background:** Low back pain (LBP) is a leading cause of work disability. While absent from work, workers with LBP may receive income support from a system such as workers’ compensation or social security. Current evidence suggests that income support systems can influence recovery from LBP, but provides little insight as to why and how these effects occur. This study examines how and in what contexts income support systems impact the healthcare quality for people with work disability and LBP and their functional capacity.

**Methods:** We performed a realist review, a type of literature review that seeks to explain how social interventions and phenomena in certain contexts generate outcomes, rather than simply whether they do. Five initial theories about the relationship between income support systems and outcomes were developed, tested, and refined by acquiring and synthesising academic literature from purposive and iterative electronic database searching. This process was supplemented with grey literature searches for policy documents and legislative summaries, and semi-structured interviews with experts in income support, healthcare and LBP.

**Results:** Income support systems influence healthcare quality through funding restrictions, healthcare provider administrative burden, and allowing employers to select providers. They also influence worker functional capacity through the level of participation and financial incentives for employers, measures to prove the validity of the worker’s LBP, and certain administrative procedures. These mechanisms are often exclusively context-dependent, and generate differing and unintended outcomes depending on features of the healthcare and income support system, as well as other contextual factors such as socioeconomic status and labour force composition.

**Discussion:** Income support systems impact the healthcare quality and functional capacity of people with work disability and LBP through context-dependent financial control, regulatory and administrative mechanisms. Research and policy design should consider how income support systems may indirectly influence workers with LBP via the workplace.

## BACKGROUND

Non-specific low back pain (LBP) is a prevalent symptom and major contributor to the global burden of disability [1-4]. Approximately 540 million people are estimated to have LBP per annum, most of whom are of working age [4, 5]. The resultant work disability leads to substantial economic burden, with the costs of healthcare and rehabilitation, wage replacement, and lost productivity cited in the billions of dollars per annum [4]. The pain, activity limitation, and participation restriction resulting from LBP can have a significant social and psychological impact, with mental health conditions a common comorbidity [6-9].

If a worker suffers from LBP that limits their ability to work they may seek wage replacement from an income support system [10]. Systems such as disability insurance, social security and workers’ compensation can provide this support. Some systems, such as workers’ compensation in Canada and Australia, fund healthcare for treatment and rehabilitation of LBP. Others, such as the Netherlands and United Kingdom (UK) fund healthcare through separate systems [10-12]. Income support systems are complex, and vary substantially between geographical regions.

Contemporary evidence regarding the management of LBP indicates that good quality healthcare adopts a biopsychosocial approach, and encourages both staying active and at work where possible [13-18]. Imaging of acute LBP and treatments such as opioids, injection therapies and surgery are considered low-value care, have high costs, and in some cases, are associated with detrimental effects on recovery and return to work [15, 19]. Healthcare quality therefore influences functional capacity, the ability of an individual to perform activities of daily living and work [19-23].

Predictors of work disability due to LBP have been characterised as falling within four domains or ‘systems’: personal, workplace, healthcare, and legislative [24]. Our review protocol hypothesised that the legislative system (i.e., income support) influences healthcare and workplace systems, which in turn impact healthcare quality and functional capacity. This hypothesis was based on contemporary literature, which suggests that income support systems may be detrimental [25-28]. For example, workers’ compensation recipients take longer to return to work than workers who do not receive workers’ compensation [29, 30].

Literature to date has typically described the relationship between income support, healthcare quality and functional capacity with a successionist model of causality. That is, X event (i.e., receipt of wage replacement from an income support system) precedes Y event (i.e., changes in functional capacity) [31], without considering the precise causal mechanism. This same literature also provides limited insight into the role of context. A greater understanding of both could improve policy design [31].

We chose realist review methodology to examine how and in what contexts income support systems influence healthcare quality and functional capacity [32]. A realist review seeks to explain how a complex social programme or phenomena generates an outcome, rather than make a determination on a causal relationship as in systematic reviews [31, 33]. This method has been applied to similar questions of complex social systems [34, 35]. In this review we sought to answer the following research questions: (1) How and in what contexts do income support systems impact the quality of healthcare in people who are unable to work due to LBP?; (2) How and in what contexts do income support systems impact the functional capacity of people who are unable to work due to LBP?

## METHODS

We conducted this review and report our findings as per Pawson’s methodology and the Realist And Meta-narrative Evidence Syntheses: Evolving Standards (RAMESES) group methodological guidance for realist reviews [36]. This involved the following stages: (1) clarifying scope, identifying review questions, formulating initial theories, (2) purposive and iterative searching for literature, (3) appraising literature and extracting data, (4) analysing and synthesising data and refining theories, and (5) disseminating review results [31, 37, 38]. In the following section we provide a summary of the research methodology. Further information on the research methods and realist theory is available in our published review protocol [32].

### Initial theories

We conducted initial purposive searches, held several collective author discussions, and consulted with experts to develop five initial theories to test in this review [32, 39, 40]. These theories were programme theories that described how and in what contexts income support systems may impact healthcare quality and functional capacity. Each theory consisted of an income support system policy that triggered a mechanism; the “non-observable” yet real process that generates an actual or empirical outcome. This mechanism acted within a set of contextual features, which modified how the mechanism generated the outcome. Initial theories were developed around the Sherbrooke Model of Work Disability [24], explaining the conceptual interactions between income support, healthcare, and workplace systems.

### Search strategy

Academic literature was the primary source of information for the refinement of our theories. Grey literature was used to define income support and healthcare system types which provided a macro-level view of policies in the regions in which studies were conducted. This provided us a greater understanding of context in each region.

#### Academic searches

We used an iterative search strategy in this review. Firstly, we conducted searches of the electronic databases Ovid MEDLINE, Cochrane Library, PsycINFO, and CINAHL. Search terms included synonyms based on the terms “low back pain”, “income support”, “workers compensation”, “social security”, “income protection”, “disability support”, “functional capacity”, “work ability”, “return to work”, “quality of care”, and “medical costs”. Search terms were combined with appropriate Boolean operators and truncations adapted to each databases’ requirements. Reference lists of included studies were also scanned and Scopus was searched to identify studies that had cited included literature.

#### Grey literature searches

Grey literature searches were conducted in the following policy databases: International Labour Organisation (ILO) NATLEX, EPLex, LEGOSH, and NORMLEX databases [41-44], United States Social Security Administration databases (US-SSA) [45], International Social Security Administration (ISSA) publication database [46], the Mutual Information System on Social Protection (MISSOC) database [47], and the Organisation for Economic Co-operation and Development (OECD) document library [48].

### Eligibility criteria

One author (MDD) screened titles and abstracts for eligibility. An additional author (TL) screened a random sample of 10% of the titles and abstracts for consistency. We included any literature that described the impact of an income support system on healthcare quality for LBP or on the functional capacity of workers with LBP. Further information on eligibility criteria is available in our protocol [32].

### Data extraction and appraisal

Data were extracted from each piece of literature by at least two authors into a standardised data extraction table within a Microsoft Excel spreadsheet [49]. As well as the characteristics of included studies (such as study region, sample characteristics and data sources), we also extracted data in relation to each of the five initial theories. The relevance and rigour of literature was rated as very, moderately, or less relevant or rigorous. Relevance refers to whether literature contains data that adequately addresses a theory, and rigour whether or not the data were generated with “credible and trustworthy” methods [31, 37, 38]. These two dimensions are typical of a realist review and are often used as a form of quality appraisal, in place of traditional quality assessment or risk of bias tools [31, 32].

### Semi-structured interviews

One author (MDD) conducted four semi-structured telephone interviews with experts in the fields of income support systems, healthcare and LBP. Interviews were not intended to be exhaustive but used to test our initial theories and identify important anecdotal or experiential knowledge that might be lacking from traditional literature searches.

Interviews were structured around each of the five initial theories; interviewees were asked whether or not they agreed with an initial theory, and why [50]. They were also asked if they thought there were any additional contextual factors or mechanisms that we had not identified. Interviews were analysed with a realist logic of analysis [50].

The Monash University Human Ethics Research Committee provided ethics approval for this project (Project ID 14144, July 2018).

### Data analysis and synthesis

Data generated from the different sources were combined and consolidated into context-mechanism-outcome (CMO) configurations [31]. These CMO configurations were first organised under each of the initial theories. Each member of the review team independently reviewed CMO configurations and theories and then the findings were discussed as a group to deliberate the role of context, the relevance and rigour of evidence, and varying outcome patterns. The review team also decided whether theoretical saturation had been reached and if subsequent further literature searches were required to adequately explain a theory. The first author (MDD) compiled the results of independent review and discussions, and refined the theories and CMO configurations. This synthesis process was performed four times, as theories were refined.

### Changes from protocol

Three minor changes were made to the methodology of the review compared to the protocol. Firstly, an eligibility criterion was added before the commencement of full-text screening: literature was excluded if it did not provide explanatory insight or understanding of how an income support system or associated policy, practice, or process impacted healthcare quality or functional capacity. This criterion was retrospectively applied to titles and abstracts from the initial search, but no additional items were included in full-text screening. The necessity of this criterion became apparent when early searches yielded large volumes of literature describing successionist causal models for the role of income support systems in worker functional capacity and healthcare quality. That is, they described simply that an interaction occurs, and did not contribute to the refinement of our theories.

Secondly, we adopted citation searching during the review (described above), as it was more efficient for iterative searches. Finally, due to time constraints we conducted four semi-structured interviews rather than the planned ten to 15 [32]. However, this was unlikely to effect the findings as the four interviews sufficiently tested the initial theories.

## RESULTS

### Search results

The first round of searching identified n=1,156 pieces of literature from academic databases (Figure 1). After duplicate removal, title and abstract and full text screening n=15 studies were included. Of the n=92 items identified from reference lists of included studies in the second round of searching, n=3 pieces of literature were ultimately included. Finally, n=520 items were identified from Scopus searches leading to an additional n=4 included pieces of literature. Once all three search rounds were completed, n=22 papers were included.

**Figure 1.**
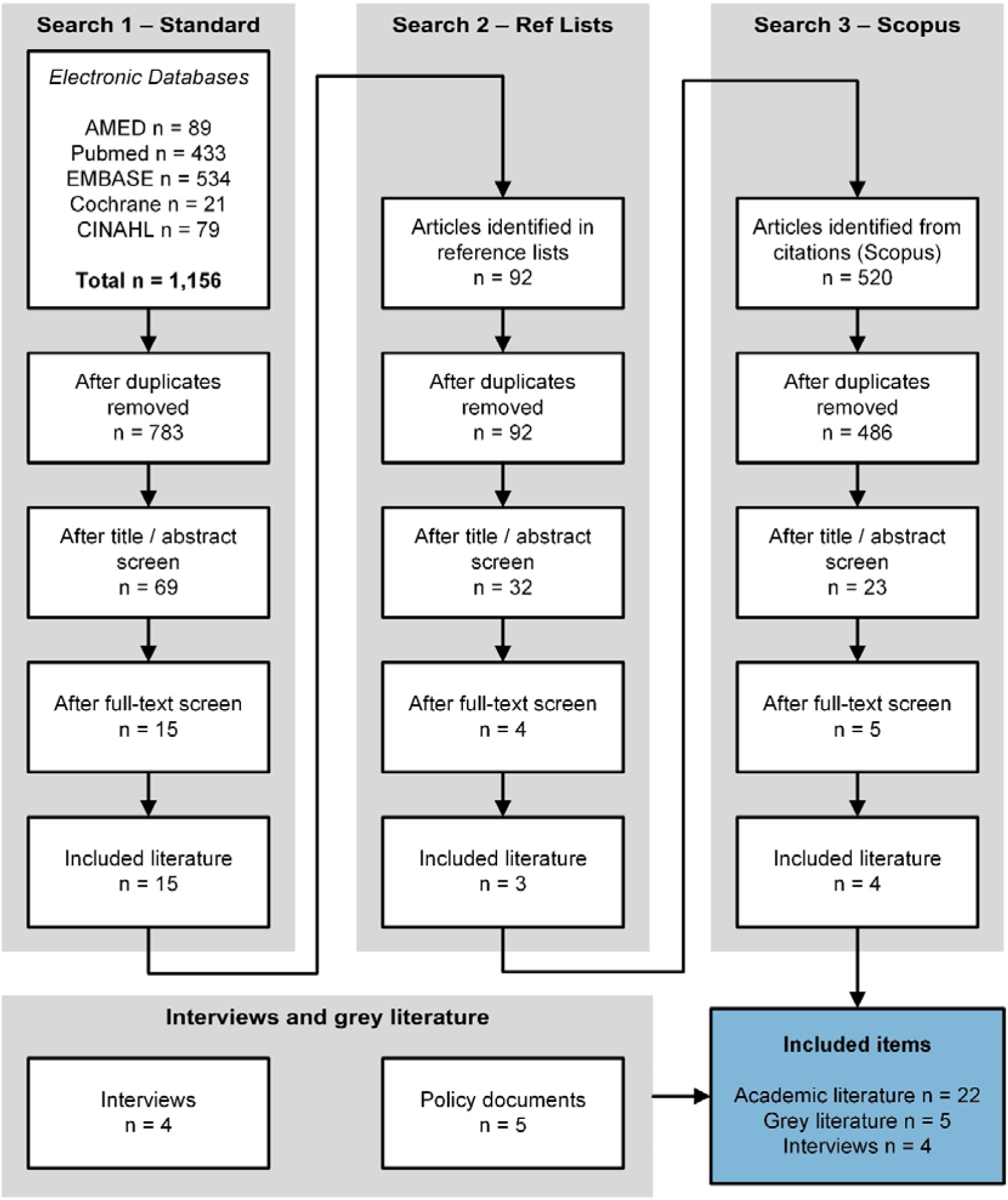
Results of searching, screening, and sorting

### Characteristics of included studies

Details of the included studies is shown in Table 1. The majority of studies were conducted in the US (n=13) and Canada (n=4). Other studies were published variously in Japan (n=1), the UK (n=1) and Australia (n=1), with a single study including six different countries (Denmark, Germany, Israel, the Netherlands, Sweden, and the US). Most studies solely utilised administrative data (n=10), six used administrative data and other data sources, five used questionnaire data alone, while two reviews used academic literature.

**Table 1.**
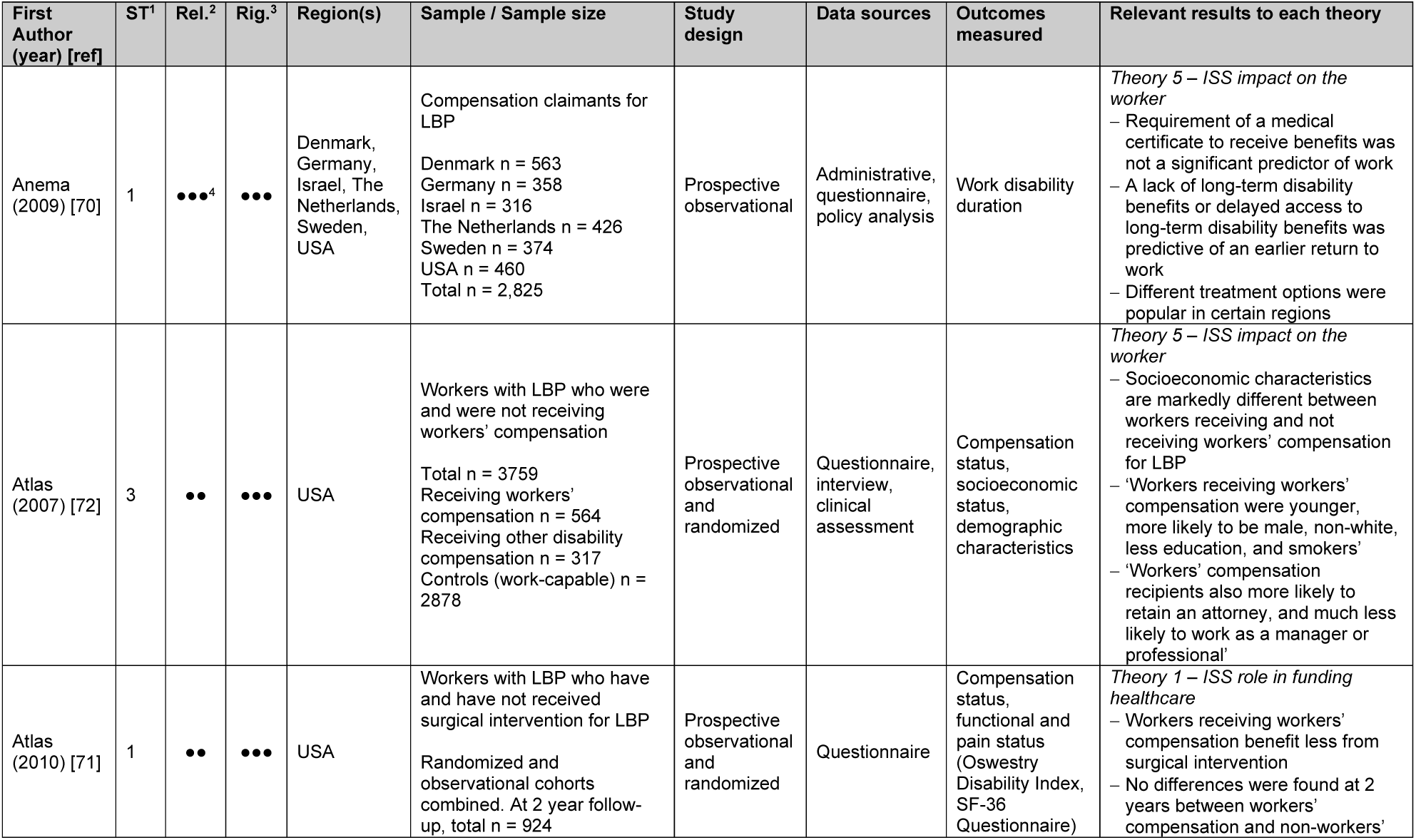
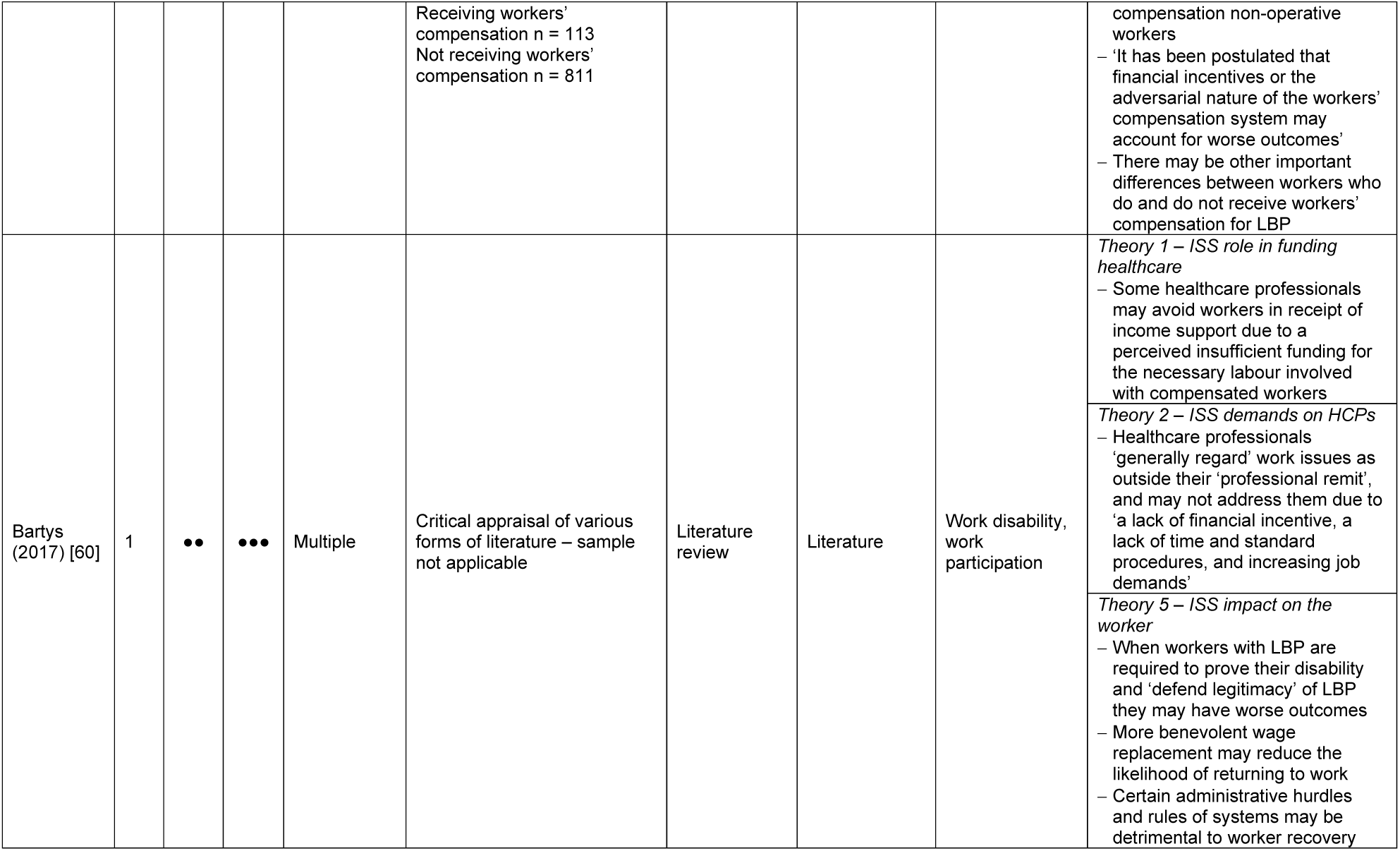
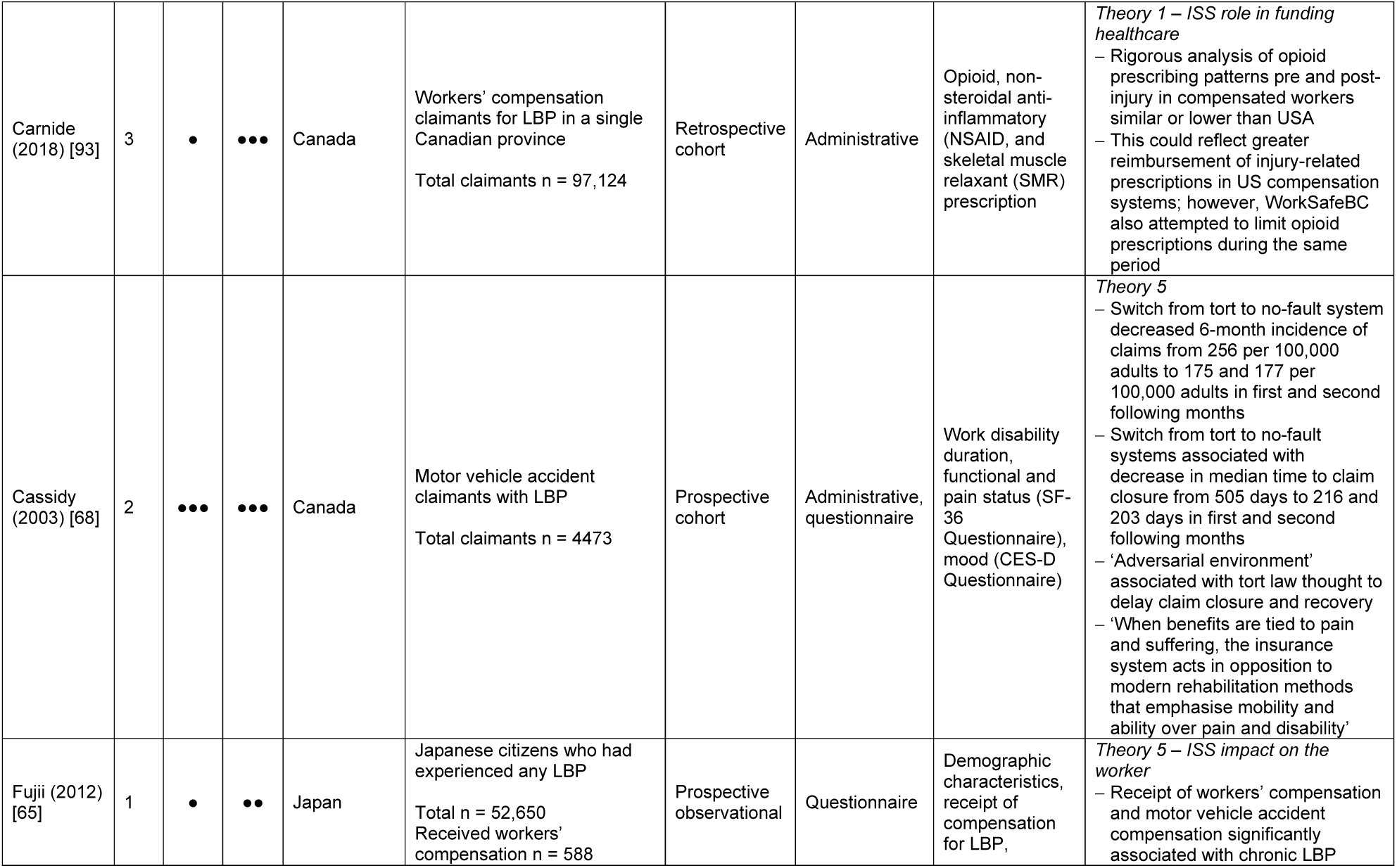
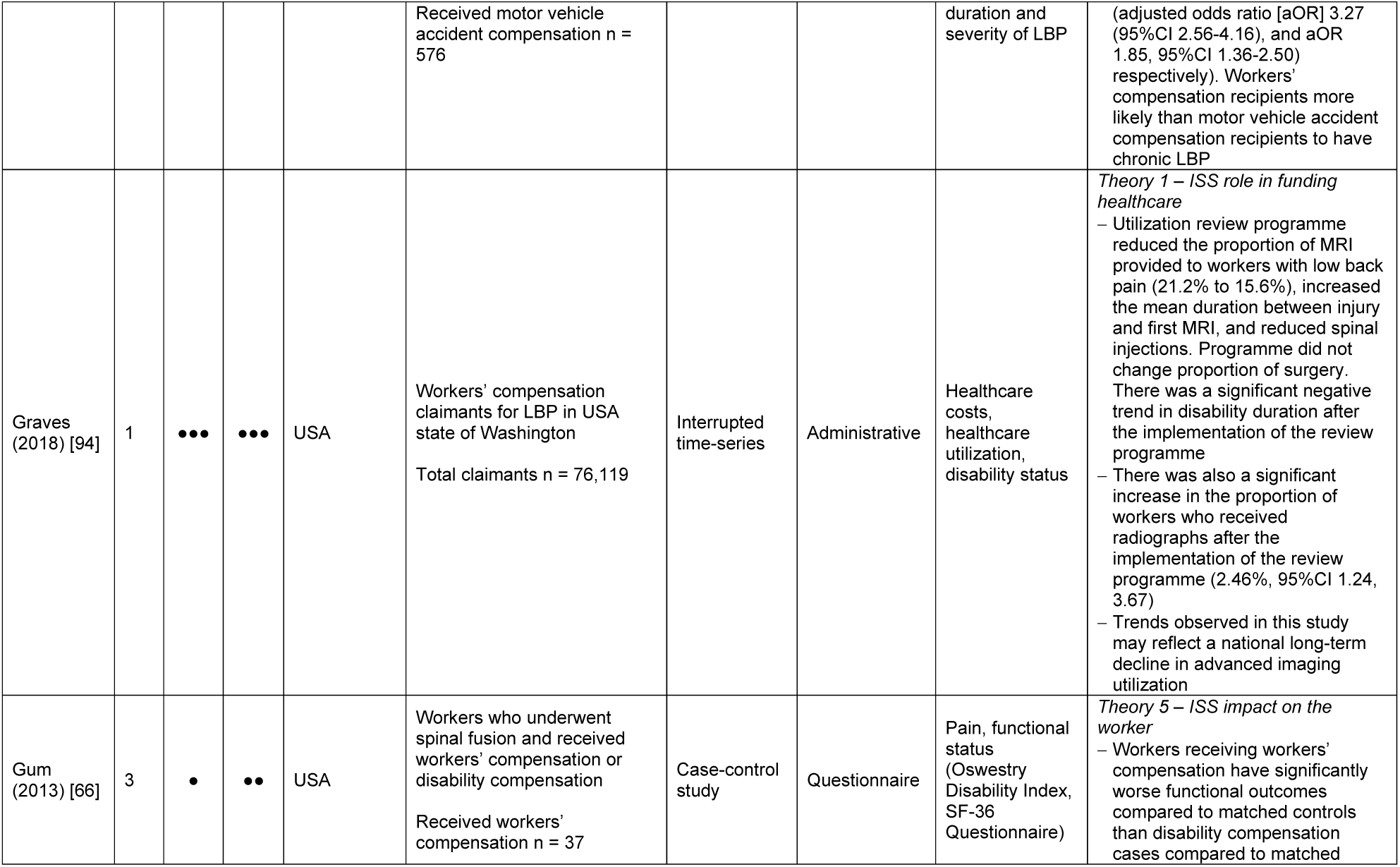
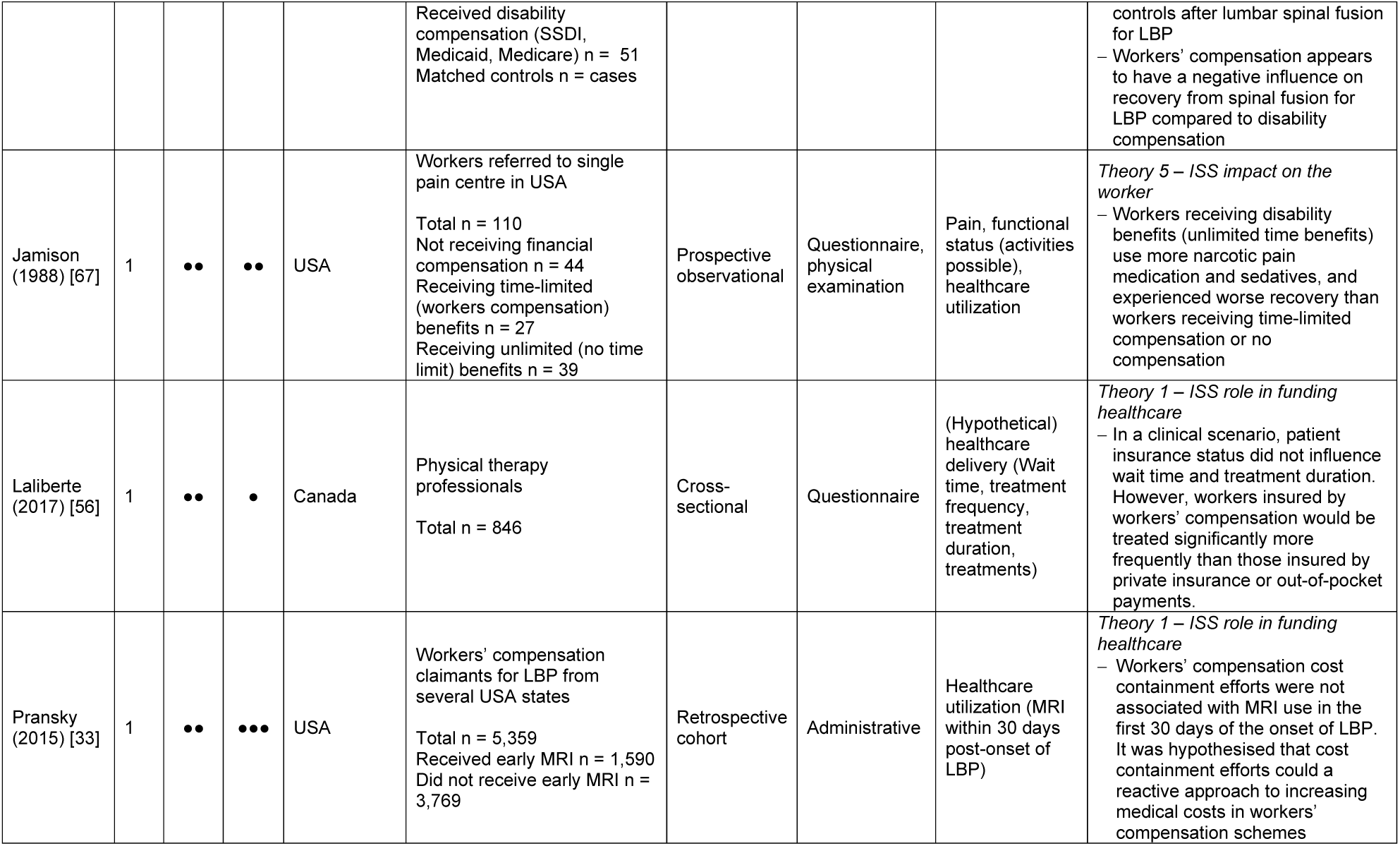
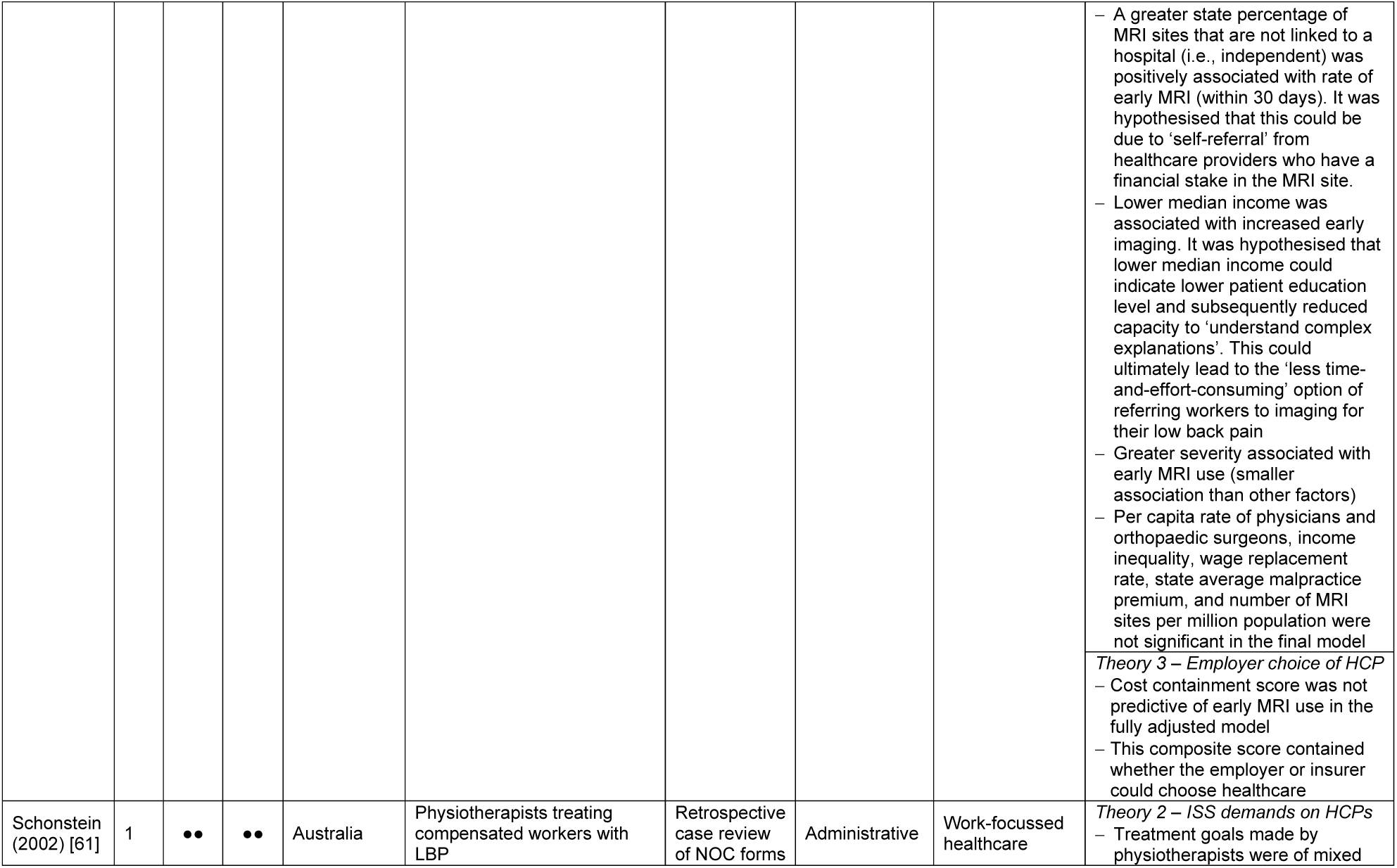
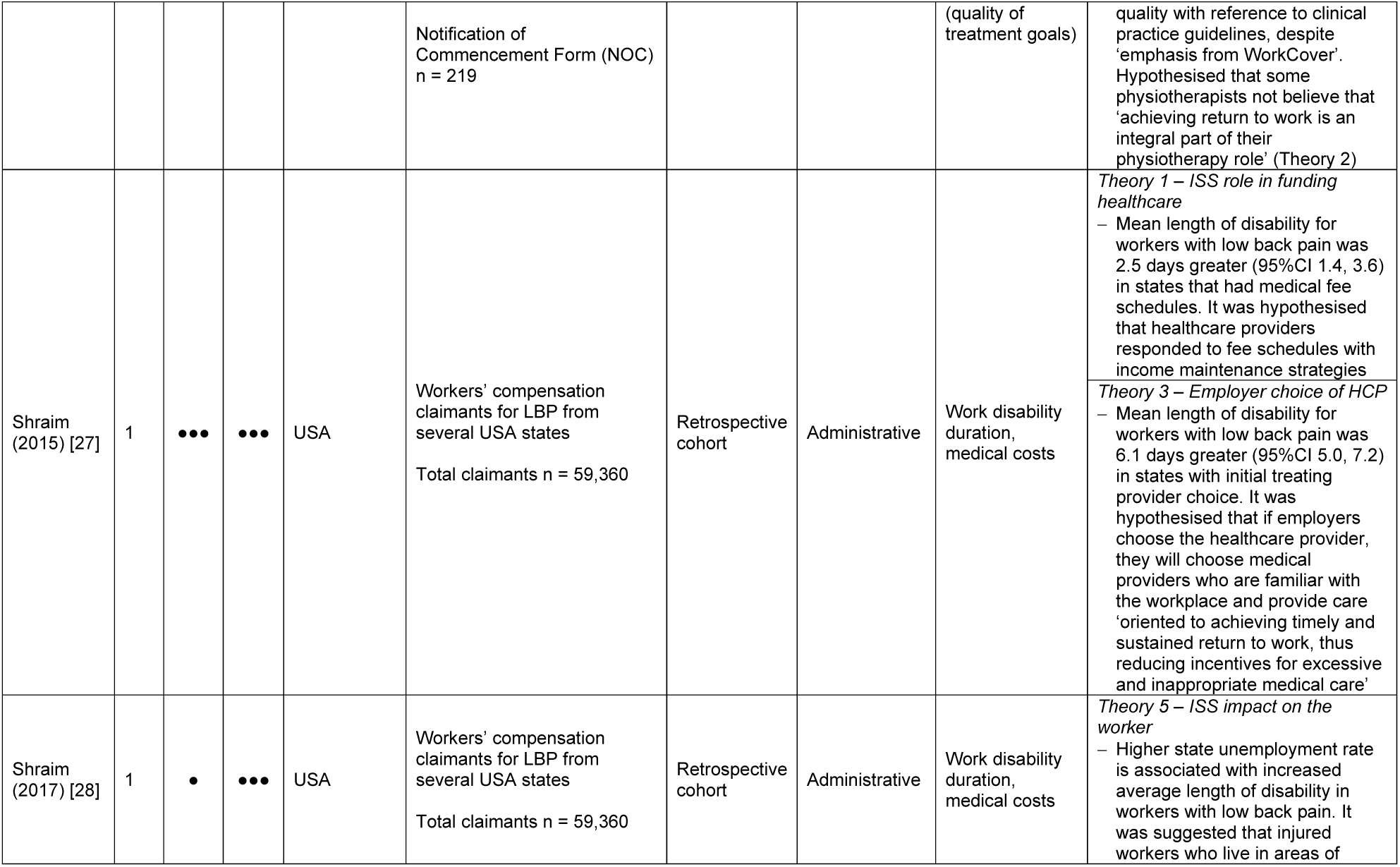
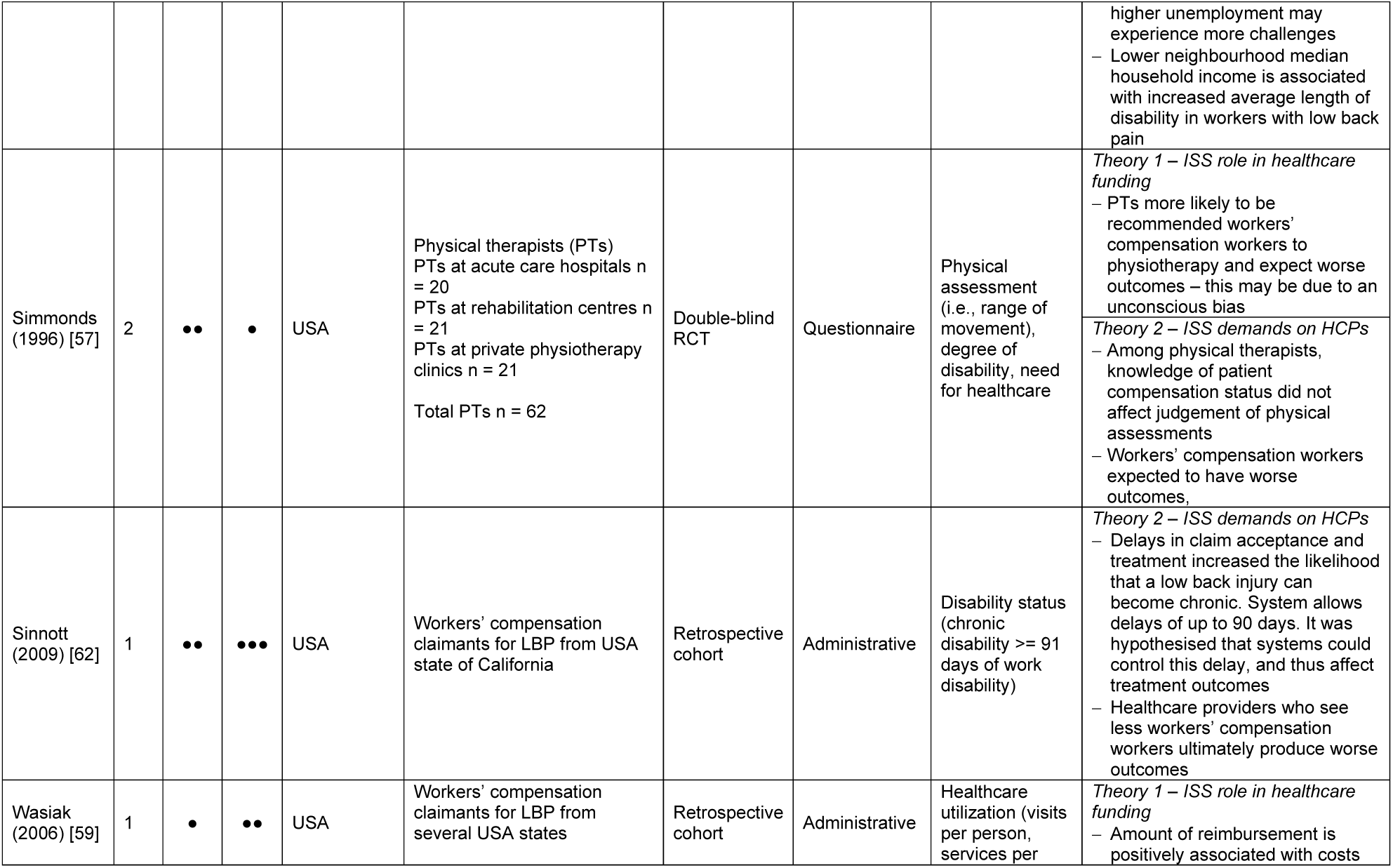
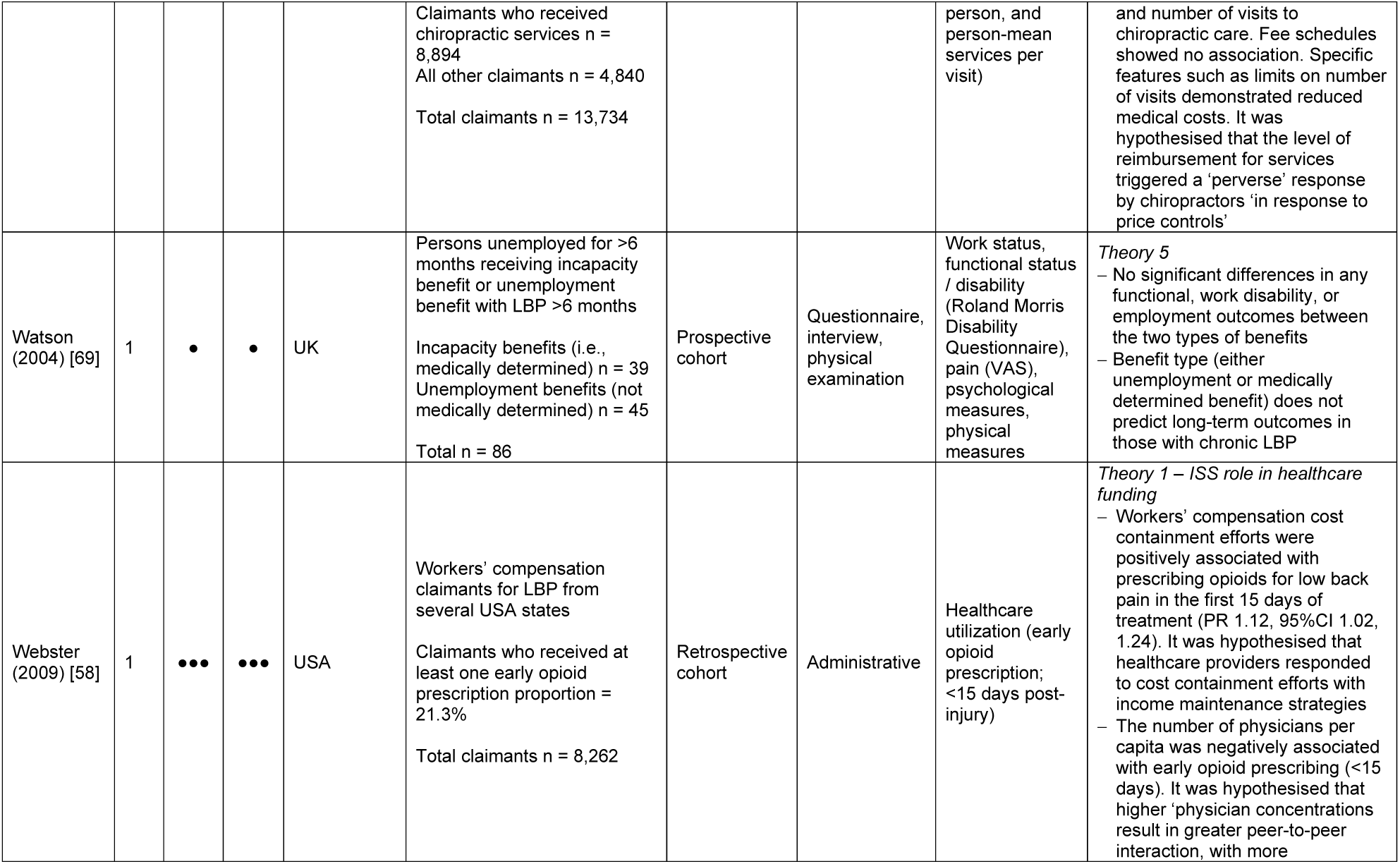
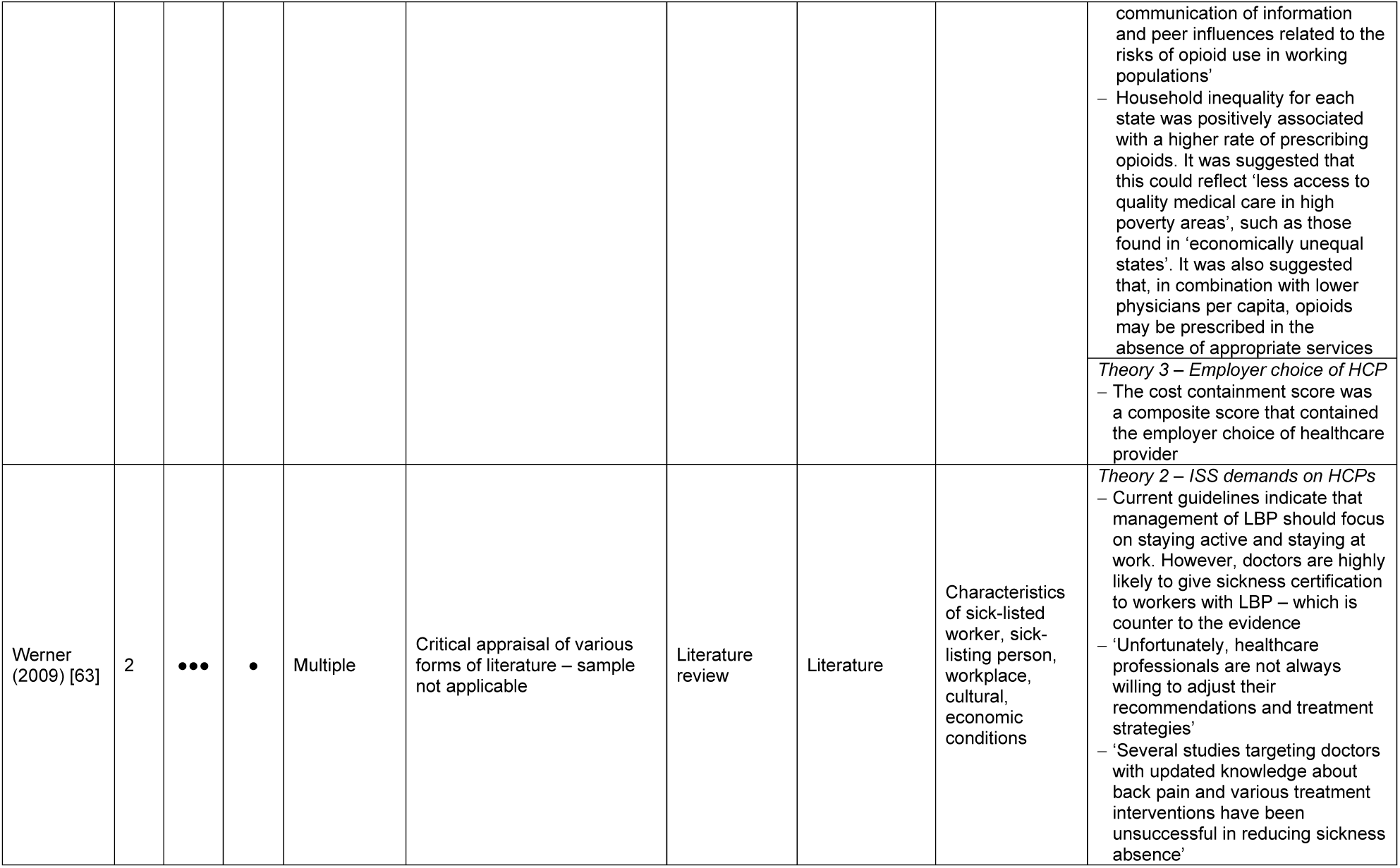
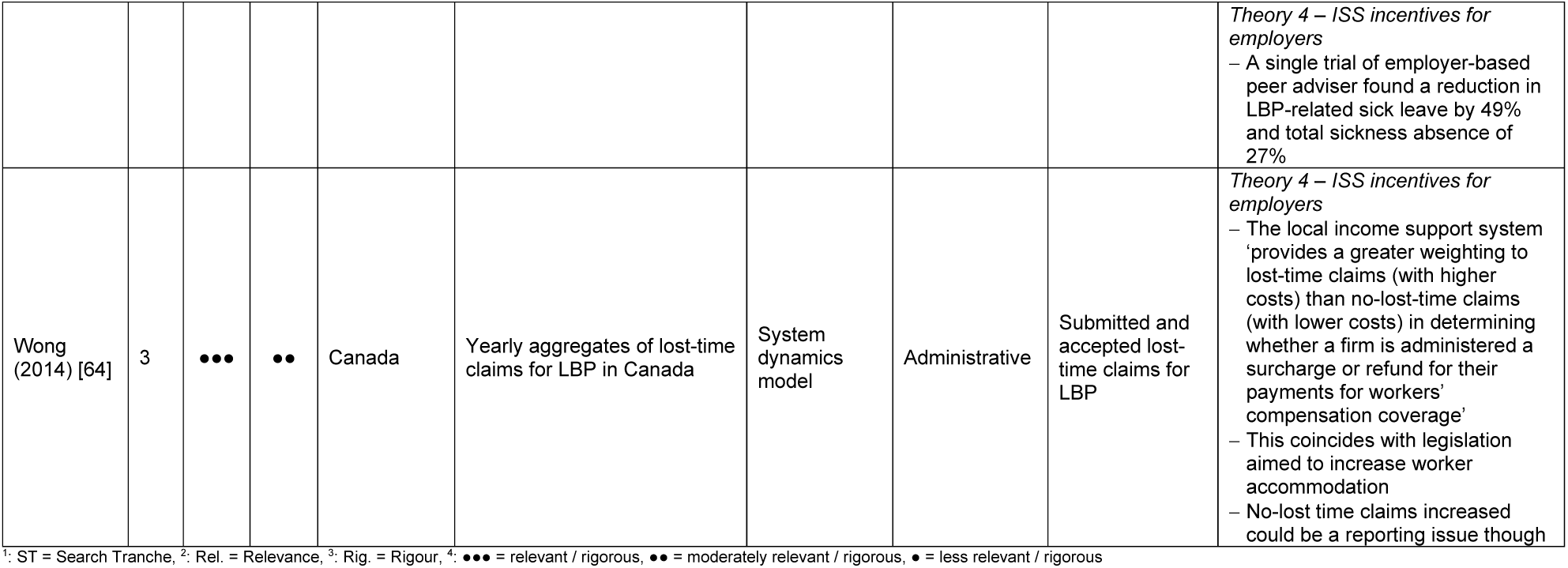
Details of included studies

### Characteristics of included systems

Information on the system types of included studies is described in Table 2. Four studies defined the typology of the disability policy model, employment injury protection scheme, unemployment protection scheme and healthcare system [51-54]. There were a limited range of system types, as most studies were conducted in North America. Most studies explored the impact of workers’ compensation systems; that is they explored the ‘employer liability’ type employment injury protection scheme. This system is a cause-based system funded by insurance premiums from employers. Three studies examined other system types, such as social security and disability insurance (SSDI). These disability-based systems sit within a liberal disability policy model and a social insurance unemployment protection scheme and benefit applicants are typically means-tested [52, 54].

**Table 2.** Characteristics and features of income support and healthcare systems in included studies [51, 53, 54]

| Country | Disability Policy Model | Employment Injury Protection Scheme | Unemployment Protection Scheme | Healthcare System Type |
| --- | --- | --- | --- | --- |
| Australia | Liberal (A) | Employer liability | Social assistance | National health insurance |
| Canada | Liberal (B) | Social insurance | Social insurance | National health insurance |
| Denmark | Social-democratic (A) | Direct provision (private insurance for accidents / public insurance for occupational disease) | Subsidized voluntary insurance and social assistance | National health service |
| Germany | Social-democratic (B) | Social insurance | Social insurance and social assistance | Social health insurance |
| Israel |  | Social insurance | Social insurance | Etatist social health insurance |
| Japan | Liberal (B) | Social insurance | Social insurance | Etatist social health insurance |
| Netherlands | Social-democratic (A) | Social insurance | Social insurance and social assistance | Etatist social health insurance |
| Sweden | Social-democratic (B) | Social insurance | Voluntary income-related insurance and social assistance | National health service |
| UK | Liberal (A) | Social insurance; social assistance | Social insurance and social assistance | National health service |
| USA | Liberal (B) | Employer liability | Social insurance and unemployment aid | Private health system |

Most studies were conducted in environments with either a private healthcare system or national health insurance system. Healthcare system type was defined by state, societal or private responsibility for regulation, funding, and service provision of healthcare. The US is one of a few systems where healthcare regulation, funding, and service provision are private. Canada and Australia have a national health insurance system in which regulation and funding are performed by the province or state respectively, with private service provision [51, 55].

Most included studies (being from North America), had income support and healthcare systems with a degree of commodification, that is, access to welfare and healthcare are dependent on market position. In the case of healthcare, this may reflect a need and desire from healthcare workers to provide services in return for income. Commodification of welfare and healthcare was not present in all studies. For example, the UK uses a national health service type healthcare system; regulation, funding, and service provision are all performed by the state leaving very little commodification.

### Interviews

Interviewed experts were from the US, Canada, UK and New Zealand and were all primarily research focussed. The sample did not include individual system actors such as employers, doctors or workers. Expert interviewee responses to initial theories highlighted the importance of context, with some providing different explanations for mechanisms compared to others. Summarised responses from expert interviewees are available in the appendix (Appendix Table 3)

### Analysis and synthesis

Results of the analysis and synthesis are presented below and summarised in Figure 2. Each theory is supported by at least one CMO configuration. Where a single CMO configuration supported a theory, the CMO configuration is reported as the theory.

**Figure 2.**
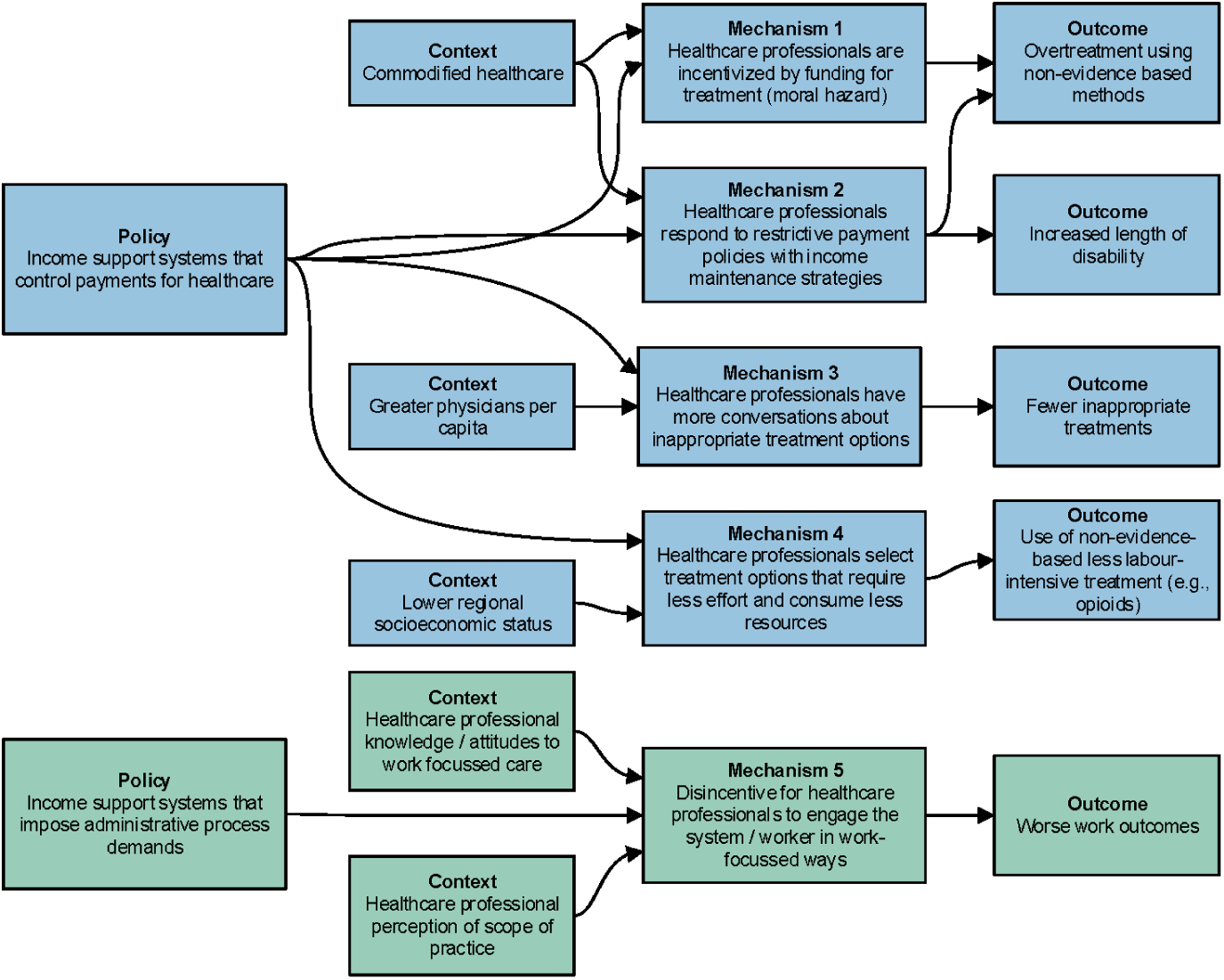
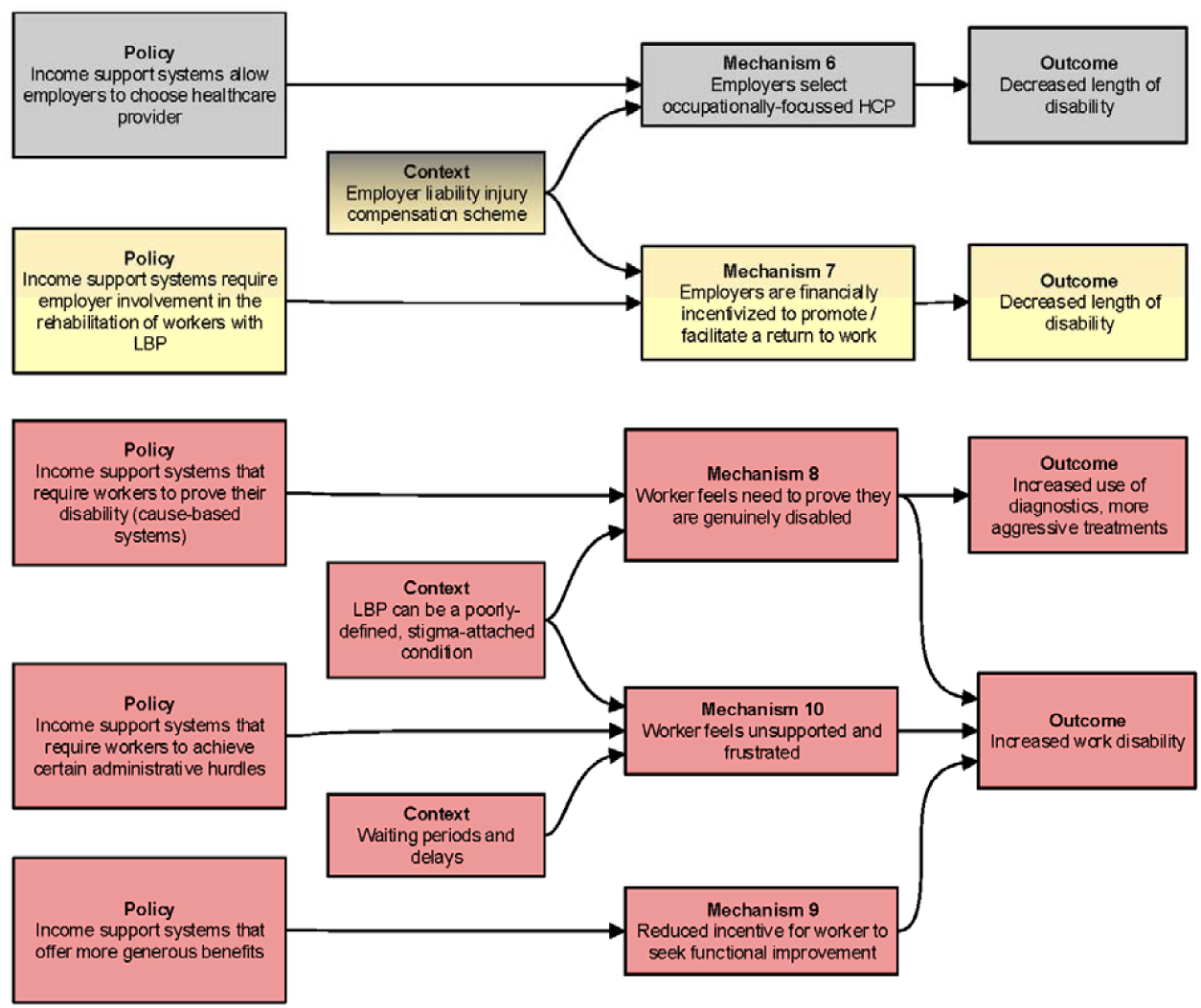
CMO configurations

### Theory 1 – Income support system role in funding healthcare

Income support systems that pay for and control healthcare in regions with commodified healthcare systems can generate adverse healthcare and functional outcomes for workers with LBP due to underlying financial incentives and healthcare professional responses to funding restrictions, as well as healthcare provider peer interactions, and regional socioeconomic factors.

#### CMO configuration 1 (n=3 pieces of literature)

*In regions where income support systems can fund healthcare that would otherwise be funded by insurance or out-of-pocket payments, healthcare professionals are incentivised to provide unnecessary care*.

Regions with a greater number of independent MRI sites had higher rates of early MRI due to potential clinician ‘self-referral’ [33]. Evidence from simulated scenarios demonstrated private clinicians are more likely to see workers with LBP more often if their healthcare was funded by an income support system (an employer liability system) than by private insurance or out of their own pocket [56]. In clinical scenarios, physiotherapists treat compensated patients similarly to other patients, but were more likely to suggest ongoing physiotherapy for treatment [57]. It was unclear whether this was due to financial incentives.

#### CMO configuration 2 (n=5 pieces of literature)

*Income support systems that attempt to control payments for healthcare in regions with private healthcare systems encourage healthcare providers to seek income maintenance strategies. This can lead to overtreatment, use of low-value treatments, and worse functional outcomes*.

Income support systems implement policy mechanisms and tools to control healthcare expenditure with strategies such as medical fee schedules, utilization review programmes, limiting treating provider choice and number of services. However, these restrictions on healthcare payments can lead to either no change in healthcare quality outcomes [33], diversion to other treatment options [20], or negative healthcare quality outcomes such as increased opioid prescribing [58], possible additional heath services [59] and poorer functional outcomes such as increased length of disability [27]. The literature suggested that healthcare providers – “increased the volume and complexity of treatment to maintain income levels” [27]. However, a fee schedule by itself does not appear to trigger a healthcare provider to demonstrate this behaviour [59]. Instead, the degree of restriction imposed by the policy tends to correlate with more income-driven behaviour [59]. This strategy was only demonstrated in studies from regions with private healthcare systems, where such a strategy is both possible to generate income.

#### CMO configuration 3 (n=1 piece of literature)

*More physicians per capita increases peer-to-peer interactions and more conversations about inappropriate treatments, leading to fewer inappropriate treatments*.

Regions with a greater number of physicians per capita demonstrated less early opioid prescribing (within 15 days) for workers with LBP [58]. A single study reported that a higher density of physicians in a given region means there is greater “peer-to-peer” interaction. It was suggested this could lead to “more communication of information and peer influences” regarding the risks associated with opioid use in workers with LBP [58].

#### CMO configuration 4 (n=3 pieces of literature)

*Healthcare providers in regions with lower socioeconomic status and fewer healthcare resources select options that require less effort, consume fewer resources and are not supported by best practice evidence*.

Regional socioeconomic factors were demonstrated to affect healthcare provision and length of disability in workers with LBP. Lower median household income was associated with higher rates of early MRI (within 30 days), as clinicians chose the “less time-and-effort-consuming” option of referring workers to imaging for LBP rather than attempting to explain the condition to workers with potentially lower education levels [33]. Household inequality was also associated with opioid prescription, as it was thought that opioids were prescribed in place of access to appropriate high-quality healthcare services in higher poverty areas [58]. Higher unemployment rates were generally associated with a greater length of disability. One offered explanation was that where unemployment is high workers have difficulty finding alternative work that is less physically demanding [28].

### Theory 2 – Income support system demands on healthcare providers

#### CMO configuration 5 (n=5 pieces of literature)

*Healthcare providers may not see work-focussed care as within their scope of practice. When income support systems impose administrative demands on healthcare providers, they lack incentives and motivation to engage the income support system and worker in a work-focussed manner, leading to poorer work outcomes*.

Work-focussed healthcare is acknowledged as important for recovery among workers with LBP among workers with LBP, yet some clinicians do not address this. Evidence from a review suggests healthcare professionals regard work issues as beyond their “professional remit” [60]. There was also evidence to suggest clinicians who do not see work issues and return to work as part of their role may focus solely on clinical issues [61]. A lack of experience with workers’ compensation workers may lead to worse worker outcomes [62], and minimal willingness to change practices based on new work-focussed knowledge [63]. Expert interviewees agreed with literature that healthcare professionals may not address work issues and income support systems because of the perceived increased demands, and lack of financial incentive, time, and decision-making authority [60]. However, a single study also suggested that knowledge of patient workers’ compensation status was unlikely to affect clinical decision-making [57]. This also aligned with statements from expert interviewees that even if healthcare providers are not incentivized to engage income support systems, quality of care was unlikely to be affected.

### Theory 3 – Employer choice of healthcare provider

#### CMO configuration 6 (n=3 pieces of literature)

*Where income support systems allow the employer to choose the workers’ healthcare provider in regions with income support systems funded by employers, the employer is incentivized by wage replacement and insurance premiums to choose the healthcare provider that will return the worker to work the fastest*.

In the US, the length of disability was lower when the employer could choose healthcare providers, which was attributed to selection of work-focussed healthcare providers who were familiar with the workplace [27]. This aligned with results from interviews. The experts suggested this theory might hold, although the employer is likely to focus on wage replacement duration as a function of total premium costs and not healthcare quality. Experts further explained that employers in their respective countries have little to no influence on healthcare. Further US literature measured the effect of limiting treatment provider choice as part of a composite cost containment score. While cost containment was shown to have an adverse effect on healthcare quality and work outcomes, it was not possible to isolate treating provider choice from the composite measure, and it is unclear how initial treating provider choice contributed to this outcome [33, 58].

### Theory 4 – Employer incentives

#### CMO configuration 7 (n=2 pieces of literature)

*Where income support systems require employer involvement in worker rehabilitation, penalties for non-compliance incentivise employers*.

There was limited LBP-specific literature available for this theory, although expert interviewees suggested that it may be possible. A single trial that found that the introduction of an employer-based peer adviser led to a 49% reduction in LBP-related sick leave [63]. Legislation to increase worker functional accommodation was followed by an increase in healthcare costs associated with no-loss-time claims [64]. Experts suggested that a financial incentive may work in their own respective systems, however the motivation would be for employers to return workers to work and reduce their financial burden, not to improve their functional capacity. Another expert pointed to a lack of financial incentive or motivation for employers to be involved in worker recovery in some regions. A third expert suggested that some employers may just ‘tick the boxes’ to meet legislative requirements until they can terminate the worker’s employment.

### Theory 5 – Income support system impact on the worker

Where income support systems with more generous benefits require workers with LBP to meet administrative requirements and prove the validity of their disability, workers feel unsupported and are not incentivized to return to work.

#### CMO configuration 8

*Income support systems that require workers to prove their injury when they have a poorly defined, stigma-attached condition such as LBP feel an ongoing need to prove they are genuinely disabled. This in turn contributes to inappropriate use of healthcare and ongoing time away from work*. (*n=5 pieces of literature*)

Where workers are required to prove they are disabled due to their LBP and ‘defend their legitimacy’, they may have worse work outcomes. It is possible that the presence of financial incentives motivate workers to have their disability recognised. Interviewed experts suggested that where workers have to prove their LBP causes disability (i.e., cause-based systems); they may seek tests that are more complex and treatments to legitimize their pain with the income support system or employer. Workers funded by workers’ compensation tended to have worse functional outcomes than workers funded by disability pensions or disability insurance (i.e., where a specific cause is not required), and motor vehicle accident insurance [65-67]. In a motor vehicle accident compensation system, a switch from a tort to no-fault scheme significantly reduced claims for LBP as well as the median duration of disability [68]. These data conflicted with a single study. However, the latter study compared types of benefits in a region with a disability-based income support system, making comparisons challenging [69]. It was hypothesised that the adversarial environment associated with tort law may delay claim closure. Experts also noted that LBP tends to carry more stigma than other ‘more credible’ conditions, and that the associated residual pain issues and diagnostic uncertainty can frustrate workers. Nevertheless, one study comparing multiple countries demonstrated that requirement for medical certification to receive benefits was not a significant predictor of engagement in return to work [70].

#### CMO configuration 9 (n=4 pieces of literature)

*Income support systems that offer more generous benefits or who step down benefits in response to return to work activities reduce the incentive for workers to seek functional improvement, contributing to increased work absence*.

Both literature and interviewed experts suggested that more ‘benevolent’ wage replacement reduced the incentive for workers to return to work [60]. Wage replacement that offered a higher proportion of the workers’ pre-injury earnings and reductions in benefits during scenarios such as partial return to work acted as disincentives to return to work. Conversely, a single trial only found differences in return to work between workers who do and do not receive workers’ compensation when they had surgical intervention for their LBP; no differences were found in those conservatively managed [71]. The authors acknowledged the potential role of financial incentives, but hypothesised that there may be other important differences between compensated and non-compensated workers. Another study of a similar cohort also identified significant differences in socioeconomic characteristics of workers who did and did not receive workers’ compensation [72]. The authors suggested these should be considered when attempting to causally link compensation and worker outcomes. An international-comparative study also suggested that the absence of long-term disability benefits, or delayed access to them, was predictive of earlier return to work [70]. The US interviewee also explained that to avoid long-term liability associated with indefinite claims in some US states, workers’ compensation bodies had begun paying lump sums to some workers, however, the effect of this was unknown.

#### CMO configuration 10 (n=1 piece of literature)

*Income support systems that include certain administrative requirements that result in waiting periods or delays and cause the worker to feel unsupported and frustrated, leading to worse functional and recovery outcomes*.

Certain ‘rules and practices’ of the system such as ‘right to case appeal’ and ‘slow or dissatisfactory case management’ have been cited as detrimental to worker recovery [60]. Interviewed experts suggested such features and waiting periods may stop the worker from working, and that if the worker feels unsupported during these periods they may be less likely to ultimately return to work. One expert also pointed to requirements for approval for certain healthcare providers or volumes of healthcare by some income support systems, possibly leading to worse functional outcomes.

## DISCUSSION

This realist review sought to understand how and in what contexts income support systems impact healthcare quality and functional capacity in workers with LBP. We have found those healthcare providers, employers, and workers’ responses to income support system policies and features, can impact healthcare quality and worker functional capacity. These effects are context specific and only operate in certain types of income support and healthcare systems or in workers with certain sociodemographic features. While contemporary literature has previously identified the impact of income support systems on these outcomes, we believe this review provides a new understanding of the causes of these events.

### Trends in included literature and theories

Several trends emerged in the included literature. The use of a successionist model of causality meant the majority of included studies treated income support systems as single entities or statuses, which may miss certain nuances inherent to large systems such as income support. The majority of included literature also did not discuss context in detail, or even at all. In particular, there was limited discussion of the context of parallel systems or regions. For example, few pieces of literature investigating income support systems also described the local healthcare system despite the potentially important relationship between the two. Finally, we found that most literature originated in regions with similar system types.

Several theoretical themes were also present. We found the prevailing mechanisms were economic incentives. The role of economic incentives in the income support and healthcare system settings has long been documented [73-77]. Most policies of income support systems rely on the responses of actors to economic incentives. In some cases economic incentives lead to contradictory behaviour from system actors. For example, the income maintenance strategy, theorised by some included studies, was thought to be a direct response to attempts to reduce healthcare costs.

### Supporting evidence from work disability literature not specific to people with LBP

We identified some evidence that was ineligible for inclusion, but may have contributed to the development and refinement of our theories. This evidence was usually excluded as the sample did specifically include workers with LBP. For the most part, work disability literature not specific to LBP aligns with our already established theories. For example, healthcare providers have previously found difficulties with workers’ compensation systems and the return to work process in workers who had “multiple injuries, gradual onset or complex illnesses, chronic pain, and mental health conditions” [26]. In some cases healthcare providers were found to refuse to treat compensated workers, citing additional “clinical complexities” and “time and financial burdens” [78]. Such evidence aligns with our established *Theory 2*. However, additional insights indicate that differences in doctors’ roles within systems may affect work outcomes [79], financial incentives may be used successfully to influence healthcare provider adoption of occupationally-focussed healthcare programs [80], and fee schedules can achieve intended cost-containment objectives [81]. Furthermore, a shift of funding income support responsibility from the state to employers was found to be beneficial for worker return to work times [82]. There is also evidence to suggest that specific financial incentives for employers may encourage actions such as claim reporting time [83].

The impact of the income support system directly on the worker is also well-documented, and aligns with our *Theory 5*. Evidence exploring the mechanisms and contexts in which benefit generosity may have incentive effects and impact work disability outcomes has previously been published [73-77]. There is also evidence to suggest that receipt of financial compensation is associated with worse functional outcomes [84]. More specifically, the administrative and legal aspects of income support systems may be generally detrimental to recovery [85], and most interactions with income support systems resulted in “significant psychosocial consequences for injured workers” [25].

However, we sought to identify and understand the impact of income support systems specifically on outcomes of workers with LBP as this is the greatest contributor to disability worldwide [4]. In the majority of cases it is not possible to identify a specific pathoanatomic cause of LBP, and the diagnosis of non-specific LBP typically depends upon ruling out much rarer specific and or serious causes. The subjective nature and causative ambiguity of LBP appears to lend itself to a certain amount of stigma or challenged integrity [86], that is not reflected in other musculoskeletal conditions such as osteoarthritis [87]. This may lead to particular challenges for a worker seeking wage replacement from a cause-based system. The US interviewee even suggested worker attempts “to prove the legitimacy of their pain and disability” might lead to greater utilisation of tests such as imaging, as reflected in *Theory 5*.

### The role of context and system types

The mechanisms identified in the review were conceptually reliant on underlying contextual factors. There is likely substantial complexity to the number and layer of contextual factors not elucidated in this review. However, we were able to understand the theoretical influence that some high-level contextual factors, such as system typology, had over some mechanisms. For example, the income maintenance behaviour demonstrated by healthcare providers appears to be reliant on a commodified healthcare system type [51, 88].

Healthcare system type is one feature of a larger policy landscape. Some included literature suggested that different income support system types (i.e., cause-based or disability-based systems) might lead to different outcomes. In one circumstance disability-based systems such as unemployment benefits appeared to have better outcomes than time-limited benefits [67]. Furthermore, it has previously been suspected that the tightening of US workers’ compensation policies lead workers to move to social security disability insurance. That is, a move from cause-based systems to a disability-based system. However, identifying causality for this inter-system movement has previously been debated [89, 90].

High level system features may also have the potential to affect how policy mechanisms in neighbouring systems react; incentives for physician performance (i.e., pay-for-performance) may function more effectively in regions within more commodified welfare systems [91]. There is potential for future research into the role of such contextual factors.

### Strengths and limitations

Realist reviews are a relatively new methodology, and provide a novel and contemporary understanding of how and in what contexts social programmes such as income support function. We believe the understanding of the role of income support systems in the population of workers with LBP benefits from the use of this methodology.

A strength of our realist review is that we used transparent methods and published our protocol including our initial theories in an open peer-reviewed journal [32]. Our protocol was peer-reviewed by two experts in realist reviews. As is customary for realist reviews we used an iterative search strategy and searched both academic and grey literature. We also documented all changes to our protocol including their justification.

We also recognise several limitations of this review. We treated context at a very high-level. By opting for system typologies, rather than more detailed features, we avoided the impact of policy changes over time and differences in local applications. However, this did prevent a more detailed understanding of the policy setting.

Included literature was published between 1988 and 2018. There has been substantial development the evidence-base for LBP, as well as in the design of income support systems, during this time. We did not account for this temporality issue in our review. Finally, we also had a small and relatively homogenous sample of expert interviewees. Although interviews were not the primary activity of this review, we could have benefited from a larger and more role-diverse sample.

### Recommendations for future research

Realist reviews are an expanding research methodology. A realist understanding can provide a nuanced understanding of the impact of social programmes and phenomena. Future research of the health and work ability of people accessing and interacting with income support systems, including those with LBP, should utilise both realist review and realist evaluation methods. We believe this will provide new insights into the role of income support systems, which have global and context-dependent ramifications.

Future research should also explore how the mechanisms identified in this review might generate outcomes in different contexts. While we are able to theorise how system typologies may enable or disable our mechanisms, we lacked the literature to explore how this might occur. A broader realist review of income support systems funding in general may be able to answer such questions. We also found that included studies rarely reported on local policies or system design. This aligns with previous research which has found that income support systems are generally defined poorly in research studies [92]. A greater understanding of context is likely to benefit future research by refining where and when policy research is generalizable.

We found a paucity of literature regarding the impact of income support system policies on the employer in cases where workers have LBP. This may be due to challenges associated with research methodologies in this cohort, or simply an under-researched area. We recommend future exploration of the impact of policies, such as financial incentives, on employers of workers with LBP.

Finally, we recommend additional policy research to examine how policies may have population-scale impact on persons and workers with LBP. This could be achieved by investigating the influence of policies in existing administrative data; a method employed by several studies included in this review.

### Recommendations for future policy development

While we acknowledge that policy development for LBP should be context dependent, we can make some broad recommendations. Firstly, a biopsychosocial approach, rather than a more typical economic approach, should be considered when developing policies.

The findings from our review confirm that there is interaction between different systems. A disabled worker typically engages a healthcare and income support system simultaneously; they require may treatment from the healthcare system to return to work, utilising wage replacement from the income support system in the interim. Any future policy development should therefore explore how policies within one system may affect other systems. We also identified research of conditions other than LBP to suggest that the policies of one income support system may affect another income support system. Future policy development should not be performed in ‘silos’, and instead consider wider societal and inter-system ramifications.

### Conclusion

This realist review has provided an understanding of how and in what contexts income support systems impact healthcare quality for and functional capacity of workers with LBP. This impact occurs through multiple underlying context-dependent mechanisms triggered by financial incentive and control policies, regulatory procedures, and administrative events. Future research would benefit from utilising realist review methods, and should make efforts to define relative contextual factors local to research. Future policy development should focus on understanding the biopsychosocial aspects of LBP, and how income support systems may interact with one another and with other systems such as healthcare.

## Data Availability

Studies included in this review are available from online digital libraries. Due to ethics agreements, notes and recordings from interviews are not publicly available.

## List of abbreviations

CMO configuration: Context-Mechanism-Outcome configuration
LBP: Low Back Pain
MRI: Magnetic Resonance Imaging
NHS: National Health Service
OECD: Organisation for Economic Co-operation and Development (OECD)
RAMESES: Realist And Meta-narrative Evidence Syntheses: Evolving Standards
SSDI: Social Security and Disability Insurance

## Declarations

### Ethics approval and consent to participate

The Monash University Human Research Ethics Committee (MUHREC) approved the semi-structured interviews performed in this review, Project ID 14144 (July 2018).

### Consent for publication

Not applicable.

### Availability of data and material

Literature included in this review is available through the relevant journals. Transcripts of interviews conducted in this review are not publicly available.

### Author contributions

All authors conceptualised the review. MDD conducted searches for academic and grey literature. MDD and TL screened literature for eligibility. MDD, TL, and RI extracted data from included literature. MDD drafted the syntheses. All authors had input on refining the syntheses. MDD drafted the manuscript. All authors provided feedback on and approved the final manuscript.

## Acknowledgements

The authors would like to acknowledge the experts who participated in the semi-structured interviews of this realist review.

## APPENDIX

**Appendix Table 3.**
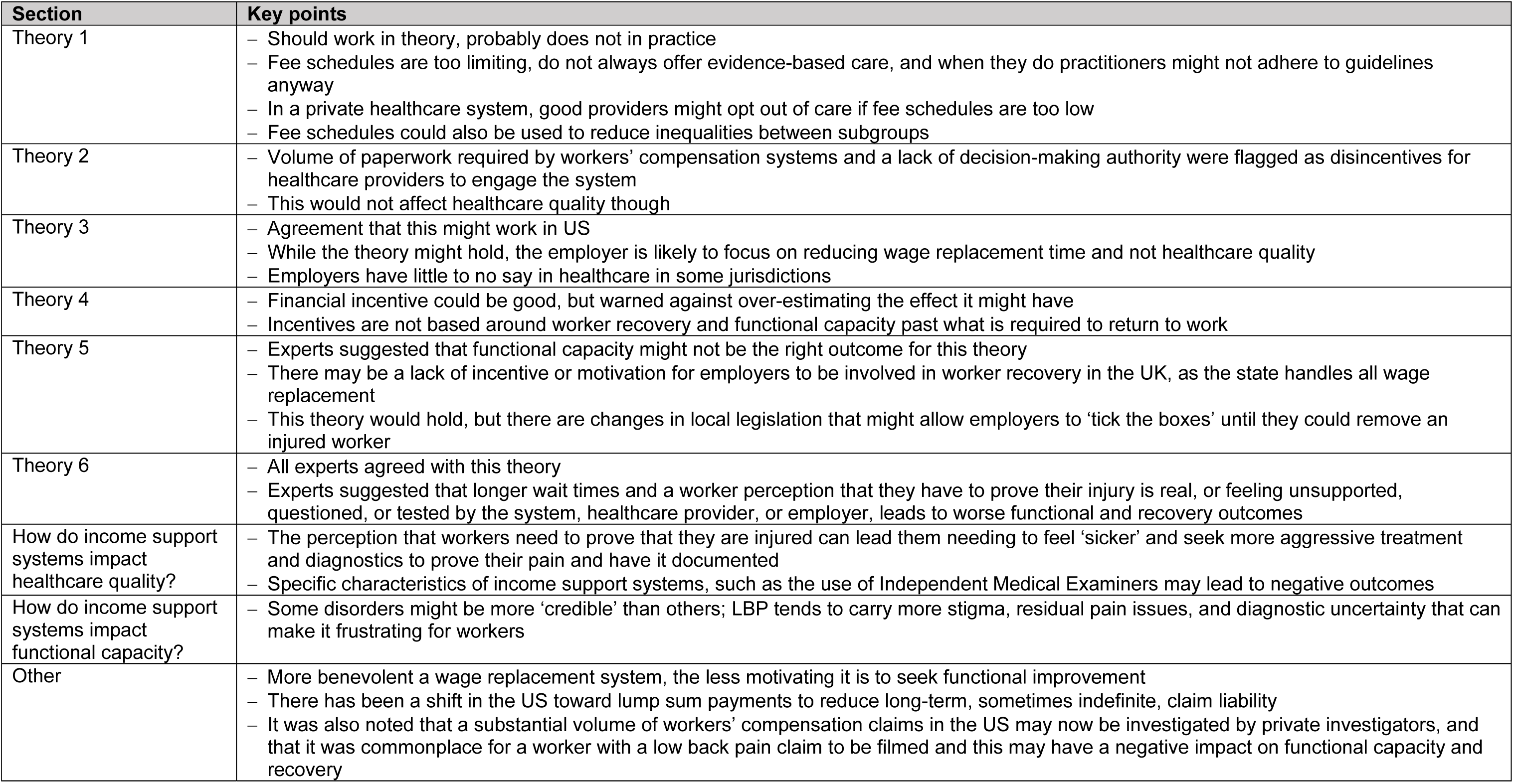
Summary of results from semi-structured interviews

